# Peripheral Inflammatory Cytokine Signature Mirrors Motor Deficits in Mucolipidosis IV

**DOI:** 10.1101/2021.03.16.21252395

**Authors:** Albert Misko, Laura Weinstock, Sitara Sankar, Amanda Furness, Yulia Grishchuk, Levi B. Wood

## Abstract

Mucolipidosis IV (MLIV) is an autosomal-recessive pediatric disease that leads to motor and cognitive deficits and loss of vision. It is caused by loss of function of the lysosomal channel transient receptor potential mucolipin-1 and is associated with an early pro-inflammatory brain phenotype, including increased cytokine expression. We thus hypothesized that peripheral blood cytokines would reflect inflammatory changes in the brain and would be linked to motor dysfunction. To test this, we collected plasma from MLIV patients and parental controls concomitantly with assessment of motor function using the Brief Assessment of Motor Function and Modified Ashworth scores. We found that MLIV patients had prominently increased cytokine levels compared to familial controls and identified profiles of cytokines correlated with motor dysfunction, including IFN-γ, IFN-α2, IL-17, IP-10. We found that IP-10 was a key differentiating factor separating MLIV cases from controls based on data from human plasma, mouse plasma, and mouse brain. Like MLIV patients, IL-17 and IP-10 were up-regulated in blood of symptomatic mice. Together, our data indicate that MLIV is characterized by increased blood cytokines, which are strongly related to underlying neurological and functional deficits in MLIV patients. Moreover, our data identify the interferon pro-inflammatory axis in both human and mouse signatures, suggesting an importance for interferon signaling in MLIV.

## Introduction

Mucolipidosis type IV (MLIV) is a neurodevelopmental and neurodegenerative disorder caused by loss of function of mucolipin 1 (TRPML1), a lysosomal channel encoded by the *MCOLN1* gene. Patients typically present with delayed developmental milestones in the first year of life and reach a plateau in psychomotor function equivalent to the 18 to 20-month range (Altarescu et al., 2002). Across their lifespan, patients exhibit progressive visual impairment due to retinal degeneration and corneal clouding, leading to blindness, achlorhydria, worsening muscular hypertonicity, and deteriorating motor function (Altarescu et al., 2002; Chitayat et al., 1991; Pradhan et al., 2002). All these features are recapitulated in the *Mcoln1* knockout mice (*Mcoln1*^*-/-*^) (Chandra et al., 2011; Grishchuk et al., 2015; Grishchuk et al., 2014; Grishchuk et al., 2016; Venugopal et al., 2007). Atypical MLIV patients with milder neurological impairment have also been recognized and the attenuated disease severity attributed to residual TRPML1 function demonstrated with some allelic variants.

MLIV results in a hypomyelinating leukodystrophy and progressive degeneration of the subcortical white matter tracts and cerebellum. Brain imaging in MLIV patients and histopathological evaluation of *Mcoln1*^*-/-*^ mice demonstrate a paucity of subcortical white matter, hypoplasia/dysgenesis of the corpus callosum, and variable white matter lesions (Frei, Patronas, Crutchfield, Altarescu, & Schiffmann, 1998). In patients, subcortical white matter and cerebellar volumes decrease with age while cortical gray matter volumes are relatively preserved (Schiffmann, Mayfield, Swift, & Nestrasil, 2014). In parallel, pathological abnormalities in *Mcoln1*^*-/-*^ mice primarily manifest in the form of reduced myelination, astrocytosis, microgliosis and partial loss of cerebellar Purkinje cells, while cortical neuron populations are largely unaffected (Grishchuk et al., 2015; Grishchuk et al., 2014; Mepyans et al., 2020).

We previously found that signs of neuroinflammation, including astrocytosis, microgliosis, and increased expression of numerous pro-inflammatory cytokines/chemokines, are present in the brains of *Mcoln1*^*-/-*^ mice early in the course of disease, before the functional symptoms first emerge (Weinstock et al., 2018). Because a growing body of data suggests that cytokine/chemokine signaling may affect the maturation and function of oligodendrocytes, (Jana & Pahan, 2005; Schmitz & Chew, 2008) Purkinje cells (Gruol & Nelson, 2005; Shim et al., 2018), and drive chronic neurodegeneration (Baune, 2015; Bosch & Kielian, 2015), we reasoned that cytokine/chemokine signaling may play a role in the pathogenesis of MLIV. Additionally, our prior work identified a marked increase in interferon gamma inducible protein 10 (IP-10) in both whole brain cortical tissues and isolated astrocytes from *Mcoln1*^*-/-*^ mice, suggesting that the interferon pathway may be strongly linked to disease progression in MLIV.

Because interferons are known to pass the blood-brain barrier (Pan, Banks, & Kastin, 1997) we set out to identify the cytokine/chemokine signature in plasma from MLIV patients and *Mcoln1*^*-/-*^ mice. We hypothesized that interferon levels would be strongly linked to motor dysfunction in human MLIV patients. To test this, we collected blood plasma from MLIV patients and familial controls while simultaneously measuring gross and fine motor scores in MLIV patients. By quantifying 41 cytokines/chemokines in the plasma, we found that cytokines were broadly up-regulated in MLIV patients compared to familial heterozygous controls. Moreover, among MLIV patients, those with reduced fine or gross motor function have robustly increased blood cytokines, including TNF-α, IFN-γ, and IFN-α2. To test if these findings would translate to mice, we also analyzed 32 cytokines in blood plasma from *Mcoln1*^*-/-*^ mice at one, two, and six months of age, corresponding to the pre-symptomatic, early-symptomatic and late stages of the disease in mice. Our analysis revealed a robust pro-inflammatory cytokine profile that changed with age, including increased IP-10 at all time points, and increased IFN-γ by 6 months of age. Finally, we found that several blood cytokines from human or mouse plasma overlapped with cytokines expressed in the MLIV mouse brain, including IP-10, suggesting that these cytokines may be candidate biomarkers of brain pathology in MLIV. In total, our findings reveal that MLIV blood cytokine signatures are strongly associated with severity of motor dysfunction in patients and suggest that peripheral cytokine signature may reflect the brain’s neuroinflammatory milieu.

## Results

### Mucolipidosis Type IV Patients Exhibit Pro-Inflammatory Blood Signatures Compared to Familial Controls

Having previously identified robust neuroinflammatory changes in the CNS of *Mcoln1*^*-/-*^ mice (Weinstock et al., 2018), here we aimed to identify a cytokine signature associated with MLIV in patient plasma compared to familial controls. We collected samples from 18 MLIV patients, including 3 mild cases (**Table 1**), and 18 parental controls and used a Luminex multiplexed immunoassay to simultaneously quantify 41 cytokines from each sample. Our analysis revealed striking differences in blood cytokine levels in MLIV patients compared with controls (**Fig. 1A**). To account for the multidimensional nature of the data, we used a discriminant partial least squares regression (D-PLSR) to identify cytokines that best distinguished MLIV patient samples from controls (Eriksson, Johansson, Kettaneh-Wold, & Wold, 2006; Weinstock et al., 2018). The D-PLSR analysis identified a weighted profile of cytokines, called a latent variable (LV1), that best distinguished MLIV samples from controls (**Fig. 1B**). Error bars representing mean±SD were generated by iteratively leaving K=5 samples out and regenerating the D-PLSR model and indicate that the model is not disproportionately influenced by a small number of samples (leave K-out cross validation, LKOCV). Scoring all the samples (**Fig. 1A**) based on the cytokine profile in LV1 separated control samples to the left and MLIV samples to the right (**Fig. 1C**). Importantly, the analysis found mild cases to group together with control cases, suggesting that the cytokine signature on LV1 segregates samples based on clinical severity (**Fig. 1D**). Univariate analysis of the top 5 cytokines from LV1 revealed significant individual differences between typical MLIV and familial control patient samples (**Fig. 1E**), but not between mild and control samples.

**Table 1:** Clinical and genotype data for study subjects.

| **Patient** | **Age**<br>**(years)** | **Mode of Diagnosis** | **Phenotype** | ***MCOLN1* Genotype** |  |
| --- | --- | --- | --- | --- | --- |
| 1 | <5 | Genetic | Typical | c.304 C>T | c.1405A>G |
| 2 | <5 | Genetic | Typical | c406 -2A>G | c.1336G>A |
| 3 | <5 | Genetic | Typical | c.1017_1020delACGG | c.877 G>A |
| 4 | 5-10 | Genetic | Typical | c.984 +1G>A | c.406 -2A>G |
| 5 | 5-10 | Genetic | Typical | c.406 -2A>G | c.964C>T |
| 6 | 5-10 | Genetic | Typical | c.694A>C | c.785T>C |
| 7 | 11-15 | Genetic | Typical | c.406 -2A>G | c.406 -2A>G |
| 8 | 11-15 | Genetic | Mild | c.406 -2A>G | c.406 -2A>G |
| 9 | 11-15 | Genetic | Typical | c.406 -2A>G | c.406 -2A>G |
| 10 | 11-15 | Genetic and Clinical | Typical | c406 -2A>G | Not identified |
| 11 | 11-15 | Genetic and Corneal bx | Typical | c406 -2A>G | Not Identified |
| 12 | 16-20 | Genetic | Typical | c.236_237ins93 | c.694A>C |
| 13 | 16-20 | Genetic | Typical | c406 -2A>G | c406 -2A>G |
| 14 | 16-20 | Genetic | Mild | c.236_237ins93 | c.1207C->T |
| 15 | 25-30 | Clinical and Gastrin | Typical | Unknown | Unknown |
| 16 | 25-30 | Genetic | Typical | c.406 -2A>G | c.406 -2A>G |
| 17 | 25-30 | Genetic | Typical | c406 -2A>G | c406 -2A>G |
| 18 | 25-30 | Genetic | Mild | c.920delT | c.1615delG |

**Figure 1.**
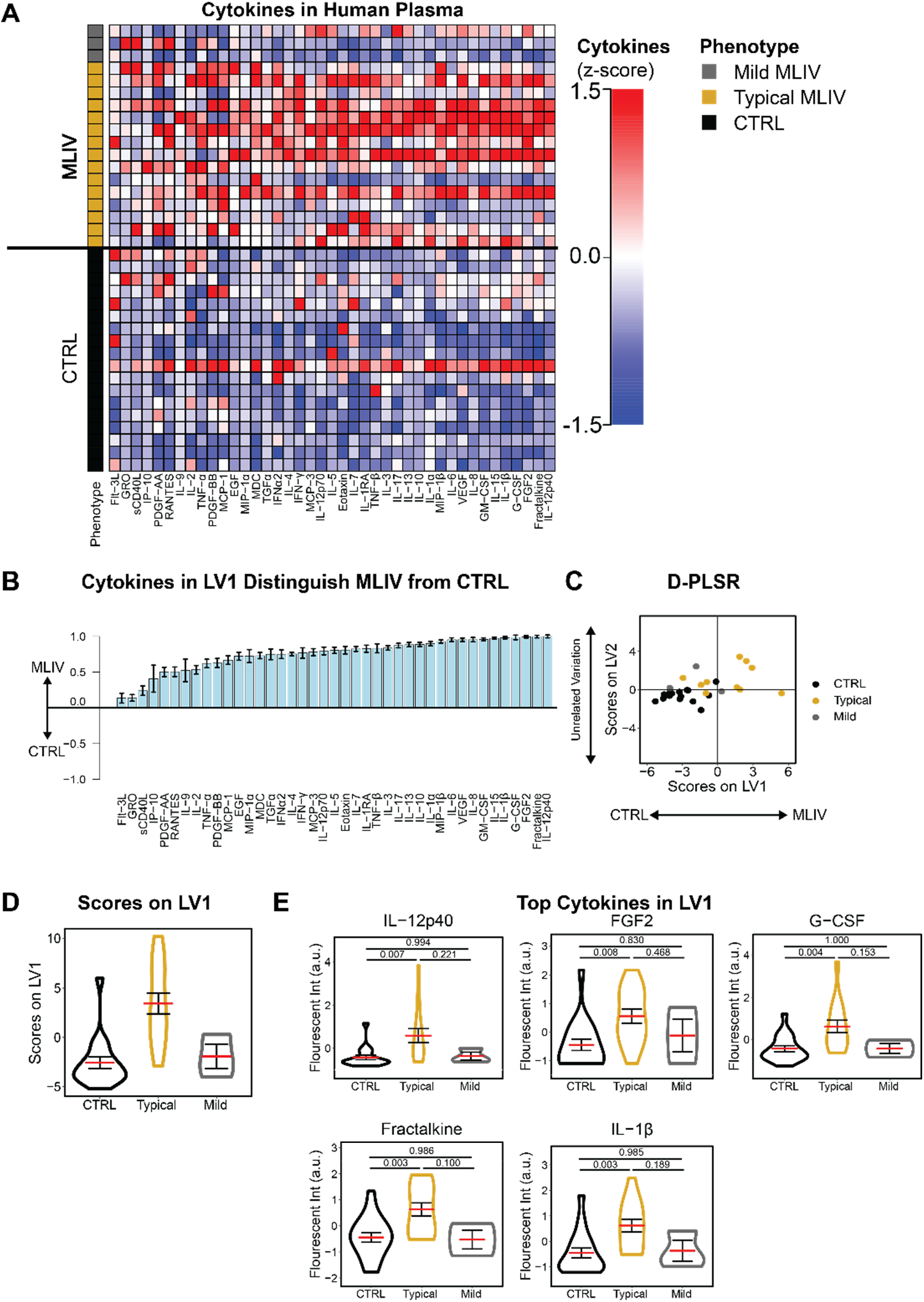
Plasma cytokines are increased in MLIV patients compared to familial controls. (**A**) Panel of 41 cytokines measured from plasma samples. Each column is z-scored. (**B**) A discriminant partial least squares regression (D-PLSR) identified a profile of cytokines, latent variable 1 (LV1) that separated MLIV cases (positive) from CTRL cases (negative) (mean±SD in a LKOCV with K=5). (**C**) LV1 separated MLIV cases to the right and both CTRL and mild cases to the right. **(D)** Scoring of each sample in (C) along LV1 (violin, mean ± SEM, N=3-15). **(E)** Univariate analysis of top cytokines from LV1, from (B) (violin, mean ± SEM, ANOVA with Tukey post-hoc correction, N=3-15).

### Plasma Cytokines are Increased in Patients with Poor Motor Function and Hypertonicity

Given the group-wise differences between typical and mild MLIV patients, we next asked if plasma cytokine levels were related to clinical disease severity. To test this, we performed analysis of plasma cytokine levels and motor function scores for each patient. We used the Brief Assessment of Motor Function (BAMF) (Cintas, Parks, Don, & Gerber, 2011; Cintas, Siegel, Furst, & Gerber, 2003; Parks, Cintas, Chaffin, & Gerber, 2007; Sonies et al., 2009) and the Modified Ashworth scale (Fosang, Galea, McCoy, Reddihough, & Story, 2003) to score domains of motor function and muscle tone, respectively, in all MLIV patients simultaneously with analysis of plasma cytokines (**Fig. 2A-B**). Interestingly, we noted that certain cytokine levels were associated with increasing age (**Fig. S1**).

**Figure 2.**
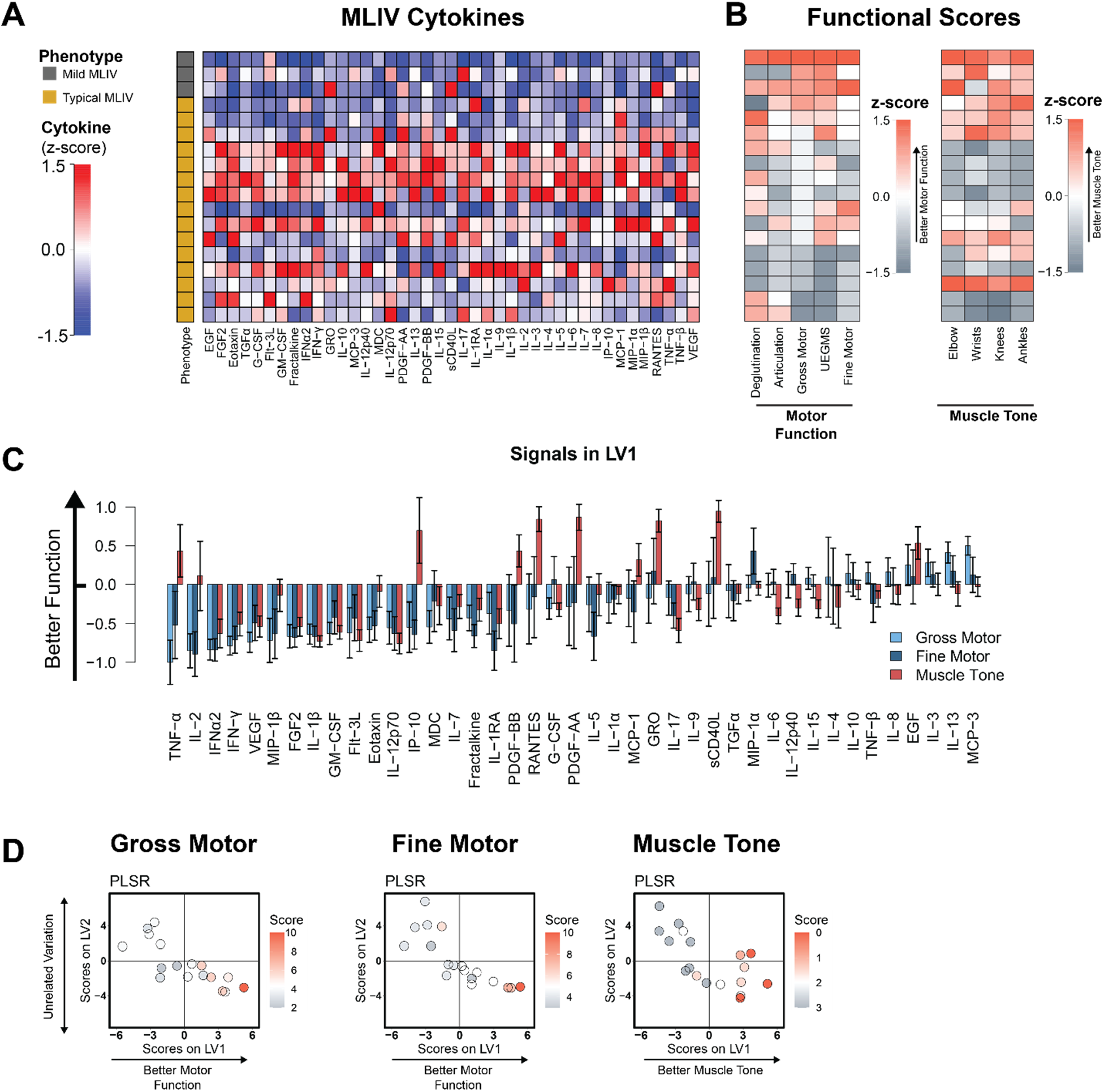
Blood cytokine signatures are related to motor dysfunction in MLIV patients. **(A)** Heatmap of relative cytokine expression for MLIV samples (columns are z-scored). **(B)** BAMF and Modified Ashworth scores for all MLIV patients (z-scored). (**C**) PLSR regressions of MLIV patient cytokines against gross motor, fine motor, and muscle tone (elbow) reveal profiles of cytokines (LV1) that correlate with poor function (negative) or better function (positive) (mean±SD in a LKOCV with K=3). (**D**) Scoring each MLIV patient sample based on the profile of cytokines for each regression separated samples with poorer function to the left and function to the right along LV1. The second axis, LV2, was associated with unrelated variation.

To identify the cytokines that were most strongly associated with the BAMF and Modified Ashworth scores, we next used partial least squares regression (PLSR) analysis to separate patients with improved motor scores to the right and with reduced motor scores to the left. Interestingly, we found that reduced gross motor and fine motor function strongly correlated with pro-inflammatory cytokines, including IL-2, TNF-α, MIP-1β, IFN-γ, and IFN-α2 (**Fig. 2C**). The PLSR analysis also revealed a profile of cytokines strongly associated with increased (i.e., worse) muscle tone. The top cytokines associated with increased muscle tone include IL-12p70, IL-1β, IFN-α2, and IL-1β. The limited overlap between cytokines correlating with lower gross/fine motor function and increased (i.e., worse) muscle tone may suggest these disease features are linked to different inflammatory processes. Importantly, we also used PLSR to correlate cytokines against both age and sex, and found that among the top 5 correlates of each, only TNF-α and IL-2 overlapped with the top 5 correlates from the gross motor, fine motor, and muscle tone analyses (**Fig. S1**). These findings indicate that the top cytokines associated with motor function or tone are only partially driven by sex and age.

### Plasma Cytokines are Increased in *Mcoln1*^*-/-*^ Mice

Having found strong relationships between cytokine profiles and motor function in human subjects, we next asked if *Mcoln1*^*-/-*^ mice exhibit a similar relationship between plasma cytokines and progression of disease. Identification of a relationship between cytokine levels and disease state in mice will enable the use of cytokines as biomarkers for pre-clinical testing. To test this relationship, we collected plasma from mice at 1, 2, and 6 months of age in male (**Fig. 3A**) and female (**Fig. S2A**) mice. We used D-PLSR analysis to identify profiles of cytokines that distinguished *Mcoln1*^*-/-*^ from wild-type controls (**Figs. 3B and S3B**). At all time-points in males, top cytokines in each profile included GM-CSF, MIP-1β, MIG, and IP-10, all of which have pro-inflammatory or chemotactic properties (**Fig. 3B-D**) (Carter, Muller, Manders, & Campbell, 2007; S. C. Lee, Liu, Brosnan, & Dickson, 1994; Peterson, Hu, Salak-Johnson, Molitor, & Chao, 1997; Vogel et al., 2015). Female *Mcoln1*^*-/-*^ mice showed a similar trend, with top correlates at 1 month of age including IL-12p70, IP-10, IL-15, MIG, all of which are chemotactic or pro-inflammatory (Carter et al., 2007; Hanisch et al., 1997; Ireland & Reiss, 2004; Jana, Dasgupta, Pal, & Pahan, 2009; Y. B. Lee, Satoh, Walker, & Kim, 1996). Interestingly, the pro-inflammatory cytokine profile was subdued in 6 month old female mice compared to controls (**Fig. S2B-D**). Together, these data demonstrate that *Mcoln1*^*-/-*^ mice exhibit a strong pro-inflammatory plasma signature. Moreover, several of these cytokines, including IP-10, are overlapping with top plasma correlates of poor motor function in MLIV patients (**Fig. 2C**), highlighting the strength of the *Mcoln1*^*-/-*^ model of MLIV for pre-clinical research.

**Figure 3:**
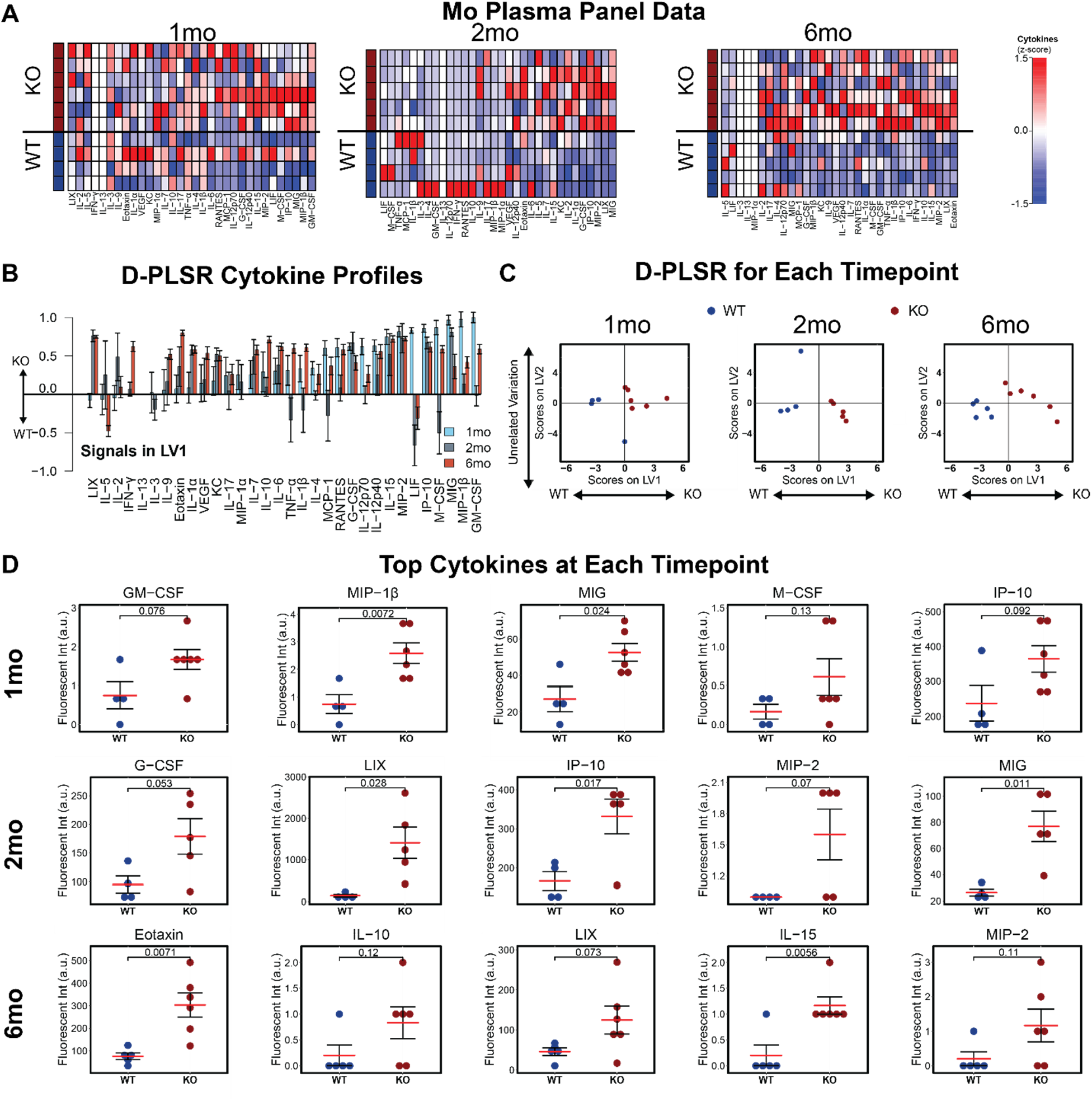
Plasma cytokine signatures distinguish male *Mcoln1*^*-/-*^ mice from wild-type controls. (**A**) A panel of 32 cytokines quantified from blood plasma in WT and *Mcoln1*^*-/-*^ animals at 1, 2, and 6 months of age (each column is z-scored). (**B**) D-PLSR analysis at each timepoint reveals cytokine signatures associated with WT (negative) or KO (positive) mice (mean±SD in a LKOCV with K=1). (**C**) Scoring each sample based on its own LV1 profile in (B) separates WT mice to the left and KO mice to the right at each time-point. (**D**) Univariate analysis of top cytokines from LV1 at each time point (mean±SEM, two-tailed t-test).

### Plasma Cytokine Signatures Overlap with Brain Cytokine Signature in *Mcoln1*^*-/-*^ Mice

A key challenge in interpreting blood cytokine signatures in MLIV patients is determining their relevance to neuroinflammation and brain pathology, given that cytokines may be sourced from multiple cell types and compartments throughout the body. Although it is impossible to fully distinguish the sources of cytokines found in the plasma, we next asked if differences in cytokines measured in the plasma from MLIV patients would reflect cytokine differences found in the MLIV brain. Since postmortem brain tissues have only been collected from one MLIV patient (Vardi, Pri-Or, Wigoda, Grishchuk, & Futerman, 2021), we instead utilized our previously published cytokine data quantified in the cerebral cortices of 2 month old female *Mcoln1*^*-/-*^ and wild-type mice (Weinstock et al., 2018) as a foundation upon which to compare our human and mouse plasma signatures (**Fig. 4A**) presented in **Figs. 1-3**. We first identified coincident cytokines that commonly separated MLIV patients from controls and *Mcoln1*^*-/-*^ from wild-type mice based on the LV1 profiles including the 26 cytokines that were overlapping between the human and mouse panels. The coincident cytokines, IP-10, IL-9, Eotaxin, IL-17, IL-13, VEGF, and IL-1α, were selected based on mean value on LV1 (≥0.2) and coefficient of variation (less than 1) (**Fig. 4B**). This reduced set of seven cytokines (i.e., coincident between human plasma and mouse brain) was able to distinguish MLIV from control samples in both human plasma and mouse brain (**Fig. 4C**). Additionally, when we used the coincident D-PLSR human plasma cytokine signature (**Fig. 4C**) to analyze mouse brain data, we found that the human plasma signature separated *Mcoln1*^*-/-*^ mouse brain samples to the right and control samples to the left. We conducted a similar analysis using the mouse plasma data to separate the mouse brain samples (**Fig. S3**). This analysis revealed five coincident cytokines between mouse brain and mouse plasma, KC, IP-10, MIG, IL-13, and IL-17, which were similarly able to separate wild-type from *Mcoln1*^*-/-*^ mouse brain samples. We noted that only IP-10 was shared between each of these coincident cytokine signatures. Given that IP-10 is a strong indicator of MLIV across human plasma, mouse plasma, and mouse brain, it is a particularly strong candidate to be considered as a predictive biomarker.

**Figure 4:**
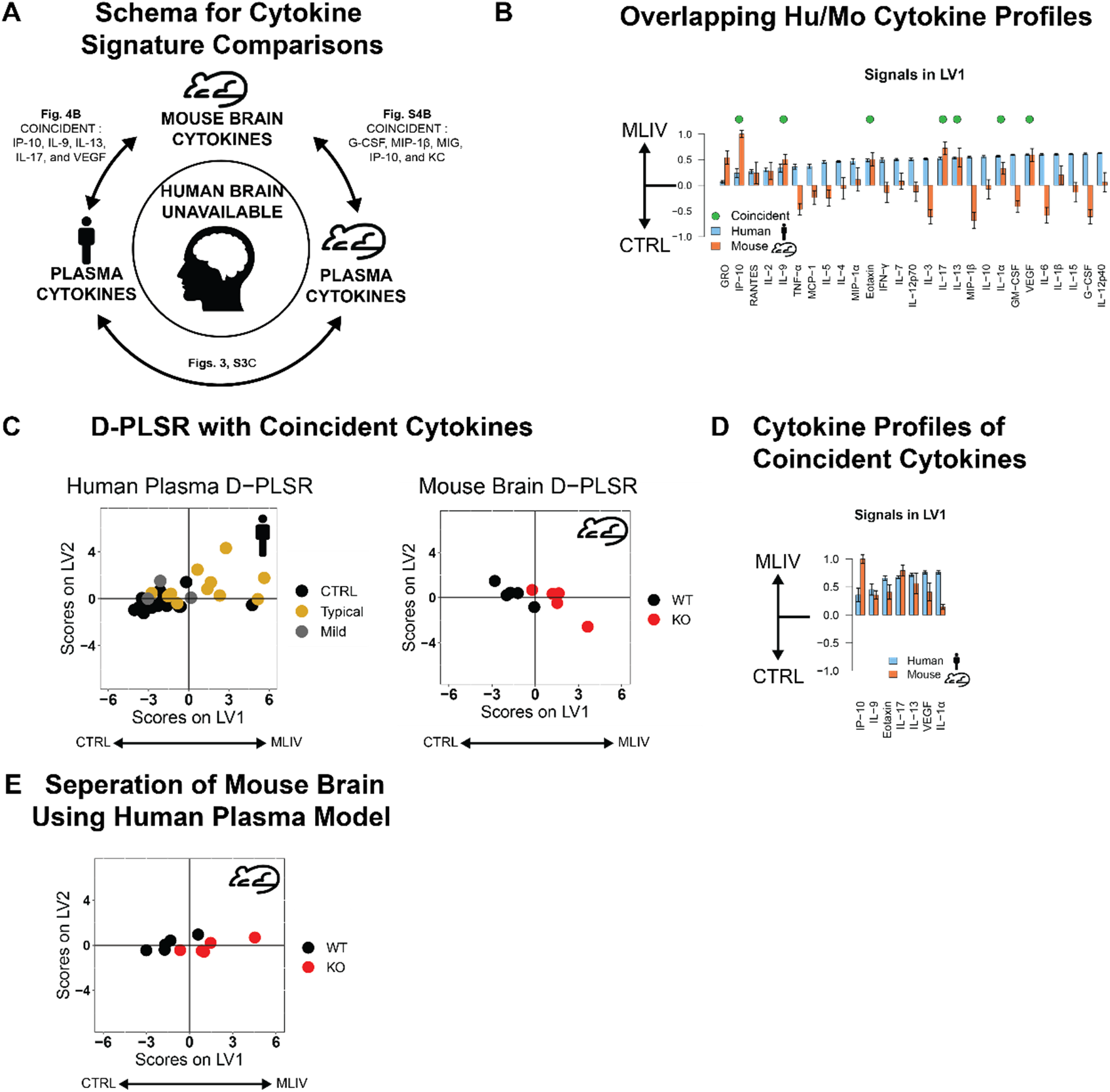
Coincident cytokine signature distinguishes human MLIV plasma and mouse brain samples. (**A**) Concept for identifying potential cytokines that may predict human MLIV brain condition based on cytokines measured in the human and mouse plasma, and on cytokine overlap between mouse brain and plasma. (**B**) LV1 cytokine profiles distinguishing mouse brain and human plasma generated based on 26 overlapping cytokines in human and mouse datasets. Of these, 7 cytokines were coincident in directionality to separate human plasma and mouse brain MLIV samples from controls (mean±SD in a LKOCV with K=3 for human samples, K=1 for mouse). (**C**) D-PLSR analysis with 7 coincident cytokines separated both human plasma and mouse brain samples from controls along LV1, suggesting that this reduced cytokine signature is capable of separating control and MLIV samples from both huma plasma and mouse brain samples. (**D**) Reduced 7 cytokine LV1 profiles distinguishing MLIV and control cases for human plasma and mouse brain. (**E**) D-PLSR model generated based on 7 cytokines from human plasma separated wild-type mice to the left and KO mice to the right based on brain cytokine measurements.

## Discussion

In this study, we extended upon our previous finding of disrupted cytokine homeostasis in the *Mcoln1*^*-/-*^ mouse brain and demonstrated profoundly altered cytokine levels in the plasma of *Mcoln1*^*-/-*^ mice and MLIV patients. Based on previous evidence of neuroinflammation in the brain of early symptomatic *Mcoln1*^*-/-*^ mice and the ability of cytokines to pass the brain-blood barrier (Banks, 2008; Brosseron, Krauthausen, Kummer, & Heneka, 2014; Pan et al., 1997; Xu, Li, & Zhong, 2015), we aimed here to establish peripheral cytokine profiles in MLIV patients and *Mcoln1*^*-/-*^ mice and test if peripheral cytokines correlate with disease progression. Indeed, our data showed that a profile of cytokines in blood plasma correlated with loss of gross and fine motor function as well as increased muscle tone. Further, we found an overlap in cytokine signatures in plasma collected from MLIV patients and *Mcoln1*^*-/-*^ mice. and between human and mouse plasma signatures and *Mcoln1*^*-/-*^ mouse brain. Taken together, our data suggest that plasma cytokine profiles reflect neuroinflammation in the CNS of MLIV patients. Further understanding of the roles of these cytokine alterations in disease pathogenesis may provide an accessible means of monitoring disease progression or suggest new targets for therapeutic intervention.

We identified pro-inflammatory cytokine signature in the plasma of MLIV patients that correlated with loss of neurological function and muscle tone. These signatures included elevations in TNF-α, IFN-α2, IFN-γ, and IP-10 (**Fig. 2C**), all of which have been implicated in microglial activation, demyelination and neuronal death (Janelsins et al., 2008; Lin & Lin, 2010; Vitner et al., 2016; Wood et al., 2015). It is important to note however, that cytokines can play both neuroprotective and neurodegenerative roles. The relationship between altered cytokine levels and their impact on the CNS is complex, and depends on the concentration, specific effects of individual cytokines, the global neuroinflammatory state, and developmental context; therefore, we cannot at present form conclusions about the mechanistic role of elevated cytokines in the pathogenesis of MLIV. However, the results of our current study lay the essential groundwork for future investigations, which could target the cytokines/chemokines identified here and test their impact on developmental myelination and/or degeneration of the subcortical white matter tracts and cerebellum.

TRPML1 has a well-established role in regulating a broad range of lysosomal functions (Colletti & Kiselyov, 2011; Di Paola, Scotto-Rosato, & Medina, 2018; Huang, Xu, Wu, Rizvi Syeda, & Dong, 2020; Wang, Zhang, Gao, & Xu, 2014) and recent data suggest its role in immune cell function (Bretou et al., 2017; Clement, Goodridge, Grimm, Patel, & Malmberg, 2020; Dayam, Saric, Shilliday, & Botelho, 2015; Goodridge et al., 2019; Santoni et al., 2020). TRPML1 activity promotes nuclear translocation of transcriptional factor EB (TFEB) (Medina et al., 2015), resulting in activation of the CLEAR network of genes, which include lysosomal hydrolases, genes of lysosomal biogenesis and autophagy (Palmieri et al., 2011; Settembre & Medina, 2015). Interestingly, transcription factors EB and E3 have also been linked to transcriptional regulation of innate and adaptive immunity, including transcription of CCL5, TNF-α and IL-1β or IL-6 (Brady, Martina, & Puertollano, 2018; Pastore et al., 2016). Therefore, the TRPML1-TFEB regulatory loop may play a central role in the transcriptional regulation of immune cells. TRPML1 has also been shown to play a role in macrophage phagophore formation, migration of dendritic cells and regulation of the effector activity of natural-killer cells Bretou et al., 2017; Clement, Goodridge, Grimm, Patel, & Malmberg, 2020; Dayam, Saric, Shilliday, & Botelho, 2015; Goodridge et al., 2019; Santoni et al., 2020). Additionally, loss of TRPML1 leads to pro-inflammatory activation of microglia and a disease-associated transcriptomic signature in the MLIV mouse microglia (Cougnoux et al., 2019), although functional consequences of these changes and role of microglia in the pathophysiology of MLIV are still not fully understood.

Though several studies demonstrate the role of TRPML1 in regulating immune cell functions in *in vitro* systems (Bretou et al., 2017; Clement et al., 2020; Dayam et al., 2015; Goodridge et al., 2019; Santoni et al., 2020), no peripheral immune-related clinical manifestations have yet been documented in MLIV patients. The reason for the lack of immune symptoms can be explained by compensatory mechanisms in immune system, potentially via an TRPML1 ortholog, TRPML2. TRPML1 and TRPML2 share structural homology and can functionally substitute each other as demonstrated in *ex vivo* studies (Curcio-Morelli et al., 2010; Venkatachalam, Hofmann, & Montell, 2006; Zeevi, Frumkin, Offen-Glasner, Kogot-Levin, & Bach, 2009). Perhaps, the most striking difference between the two channels is their tissue specificity. Unlike TRPML1, which is ubiquitously expressed at a stable level in all examined tissues and cell types, expression of TRPML2 is limited to lymphoid tissue, particularly thymus, spleen and immune cells (Cuajungco, Silva, Habibi, & Valadez, 2016; Samie et al., 2009). Therefore, it is tempting to speculate that expression of TRPML2 in this selected set of tissues may be able to prevent gross consequences of TRPML1 loss of function and spare some aspects of immune function in patients with MLIV. This is supported by data showing that TRPML1-deficient B-lymphocytes expressing TRPML2 do not manifest lysosomal pathology typically observed in TRPML1-deficient cell types (skin fibroblasts, neural cells, etc.) that do not endogenously express TRPML2 (Song, Dayalu, Matthews, & Scharenberg, 2006).The role of lysosomes in mediating multiple aspects of immune cell function is well established (Ma, Galluzzi, Zitvogel, & Kroemer, 2013; Reinheckel, 2013; Watts, 2012), and signs of neuroinflammation, including altered cytokine levels have been reported in several lysosomal storage diseases (LSD), including Gaucher disease, Fabry disease, Neuman-Pick disease and MPS (Grabowski, 2017; Rigante, Cipolla, Basile, Gulli, & Savastano, 2017; Rozenfeld & Feriozzi, 2017; Vitner, Futerman, & Platt, 2015). However, surprisingly little is known about systemic immune dysfunction in other LSDs, and a systematic approach to establish peripheral signature of cytokines/chemokines for LSDs is lacking. Here we show that blood cytokines can be linked to severity of CNS dysfunction in the lysosomal disease mucolipidosis IV, which offers new possibilities to track disease progression in MLIV patients and pre-clinical model.

Overexpression of cytokines, including interferons, in the brain during neurodegenerative process, has led to studies determining whether interferons and other cytokines can pass the blood brain barrier (Banks, 2008; Pan et al., 1997), and whether cytokines may serve as a blood biomarkers of neuroinflammatory dysfunction (Xu et al., 2015) or neurodegeneration, e.g., in Alzheimer’s disease (Brosseron et al., 2014; Chang, Wu, Chen, & Chen, 2015).

Collectively, our mouse and human data suggest an intricate relationship among neuroinflammation, peripheral cytokines, and loss of motor function in MLIV. Although our data do not reveal whether or not the commonalities between the brain and periphery are due to cytokines passing the blood-brain barrier, or a common mechanism between the brain and peripheral compartments, they do reveal that the peripheral cytokine signature is strongly associated with the motor dysfunction in MLIV. Given this robust relationship and the likelihood that new therapies will simultaneously target brain and peripheral compartments, cytokines hold strong potential to serve as an early indication of patient response to forthcoming therapeutic strategies for this devastating disease.

## Methods

### Study Design and Population

Patients with MLIV (N=18, F=9 and M=9) and familial controls (N=18, F=11 and M=7) were recruited through the Mucolipidosis Type 4 Foundation. Written informed consent was obtained from legal guardians for participation in our approved natural history and biomarkers studies according to protocols approved by the Massachusetts General Hospital Institutional Review Board. Written informed consent or assent were obtained from patients if their neurological capacity allowed. Inclusion criteria for patients included a documented diagnosis of MLIV by 1) clinical or research-based sequencing of *MCOLN1* and identification of two pathological *MCOLN1* alleles or 2) presence of the expected constellation of clinical symptoms associated with MLIV and documentation of at least one of the following: one pathological *MCOLN1* allele, elevated gastrin levels, or a tissue biopsy with evidence of lysosomal inclusions consistent with MLIV. One familial control was excluded from cytokine analysis when it was later revealed that they had undergone a surgical procedure in the week prior to sample collection which could affect cytokine levels. MLIV cases were assigned to either typical (N=15) or mild (N=3) presentation. Based on our clinical experience, we classified patients as mild if they had obtained independent ambulation at some point in life. All patients underwent a full examination by a single board-certified pediatric neurologist and their function scored with the Brief Assessment of Motor Function (BAMF) scales (gross motor, upper extremity gross motor, fine motor, deglutition and articulation) and Modified Ashworth scale. Due to limitations associated with patient access and cost, repeat scoring by independent evaluators was not possible.

### Blood Sample Collection and Processing

Blood was collected K2EDTA-coated Purple/Lavender top vacutainers (BD 368047). Plasma was isolated within 4 hours from collected blood samples by centrifugation at 1000g for 10 min at 4^0^C. Isolated plasma was stored at −80°C.

### Animal Studies and Sample Collection

*Mcoln1* knock-out mice (on a C57Bl/6J background) were maintained and genotyped as described previously (Venugopal et al., 2007). Blood was collected from *Mcoln1*^−*/*−^ and littermate *Mcoln1*^*+/+*^controls at 1, 2 and 6 months of age and plasma was isolated within 4 hours from collected blood samples by centrifugation at 1000g for 10 min at 4^0^C. Isolated plasma was stored at −80°C. All experiments were performed according to the US National Institute of Health guidelines and approved by the Massachusetts General Hospital Institutional Animal Care and Use Committee.

### Cytokine Luminex Immunoassays

Plasma was stored at −80°C. For human cytokine analysis, samples were diluted to 8% in Milliplex assay buffer and analyzed using the Milliplex MAP Human Cytokine/Chemokine Magnetic Bead Panel - Premixed 41 Plex kit (Millipore Sigma, St. Louis, MO, USA, HCYTMAG-60K-PX41). Mouse plasma was diluted to 70% in assay buffer and analyzed using the Milliplex MAP Mouse Cytokine/Chemokine Multiplex assay (Millipore Sigma, St. Louis, MO, USA, MCYTMAG-70K-PX32). Assays were read out with a MAGPIX Luminex instrument (Luminex, Austin, TX, USA).

### Correlation and Multivariate Analyses

(D-)PLSRs were performed in R (R Foundation for Statistical Computing, Vienna, Austria) using the ropls package v1.4.2. The data were z-scored before input into the function. Cytokine measurements were used as the independent variables, and the discrete regression variable in all analyses was genotype/phenotype. Orthogonal rotations were applied to the sample scores and analyte weightings to obtain consistent separation of each group along the LV1 and LV2 axes. Error bars for LV loadings were calculated by iteratively excluding K samples without replacement 100 times (leave-K-out-cross validation, LKOCV), and regenerating the D-PLSR model each time. Error bars in the LV1 plots report the mean and SD computed across the models generated to provide an indication of the variability within each cytokine among models generated. Sample outliers were checked for and removed in the mouse cytokine data by performing a principle component analysis on the data and iteratively removed data points that fell outside of a 99.5% confidence ellipse (mahalanobisQC in ClassDiscovery package v3.3.13).

## Data Availability

De-identified data are available upon request.

## Author Contributions

Y.G., A.M., and L.B.W designed and oversaw execution of the study. A.M. conducted motor function and clinical assessment of motor function scores. S.B.S, A.F and L.D.W. collected blood plasma data. L.B.W and L.D.W designed and executed computational analyses. A.M, Y.G., and L.B.W drafted the manuscript. All authors reviewed the manuscript. A.M. was selected to appear first due to his substantial writing and critical revision of the text.

## Acknowledgments

We thank the ML4 foundation, Dr. Rebecca Oberman and Randy Gold for providing research funding (Y.G) and help with blood samples collection. This work was also funded by the Upenn Orphan Disease Million Dollar Bike Run grant (to A.M. 2017D007934), NINDS/NIH (to A.M. 01K12NS098482-01) and startup funds from the Woodruff School of Mechanical Engineering at Georgia Tech (L.B.W.). L.D.W. was supported in part by the National Institutes of Health Cell and Tissue Engineering Bio-technology Training Grant (T32-GM008433). The authors are also grateful to Raffi Shchiffman (Baylor Scott & White Research Institute

Dallas, Texas) and Annick Raas-Rothschild (Department of Human Genetics, Hadassah Hebrew University Hospital, Israel) for valuable discussions on this work and to Alysa Pybus for assistance with computational analysis.

