## Supplementary Information for "Peripheral Inflammatory Cytokine Signature Mirrors Motor Deficits in Mucolipidosis IV"

### Supplementary Figures

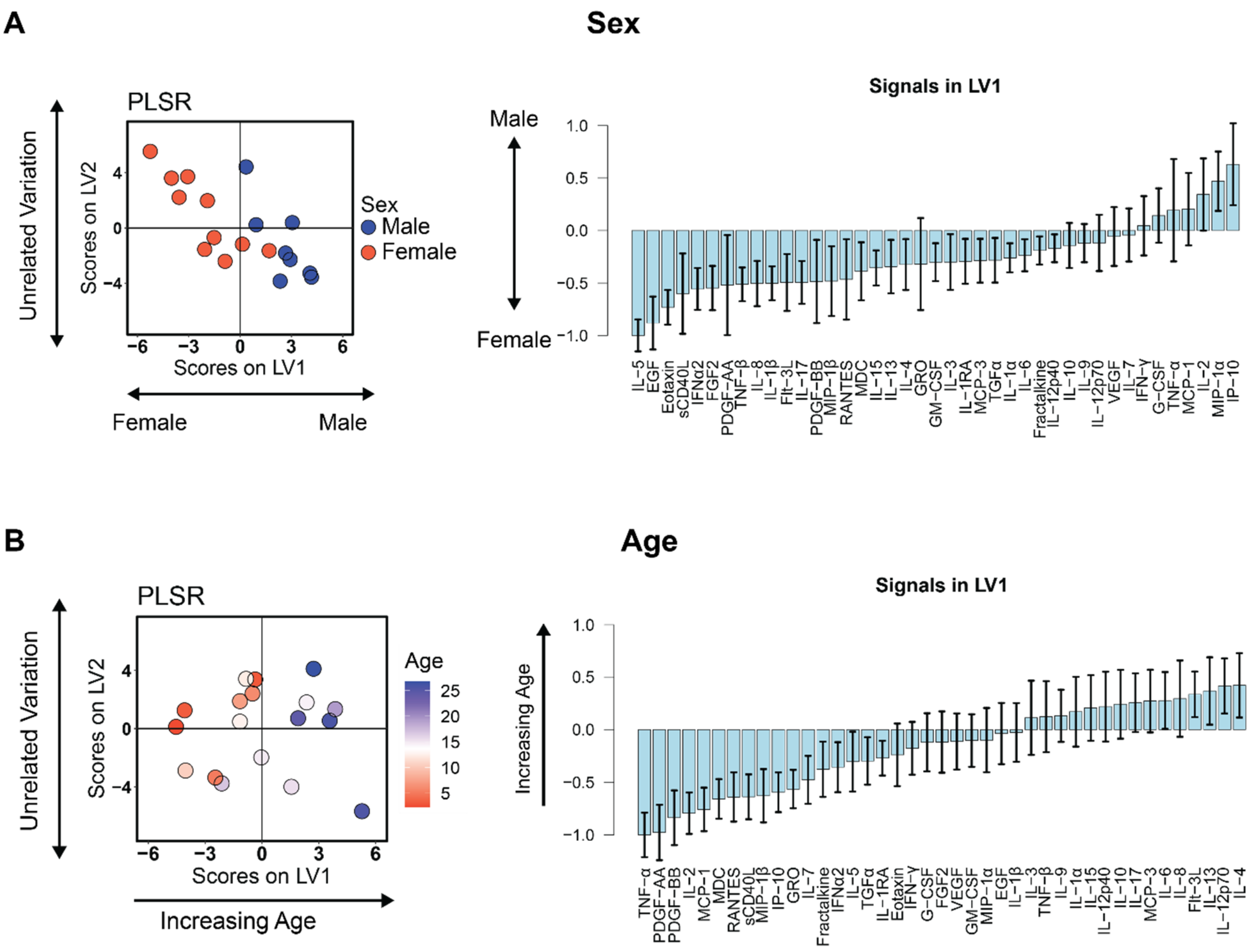


**Figure S1: Blood cytokine signatures are related to sex and age in MLIV patients.** (**A**) PLSR regression of MLIV patient cytokines against sex revealed a profile of cytokines (LV1) that correlate with females (negative) or males (positive) (mean±SD in a LKOCV with K=3). (**B**) PLSR regression of MLIV patient cytokines against age revealed a profile of cytokines (LV1) that correlate with increasing age (mean±SD in a LKOCV with K=3).

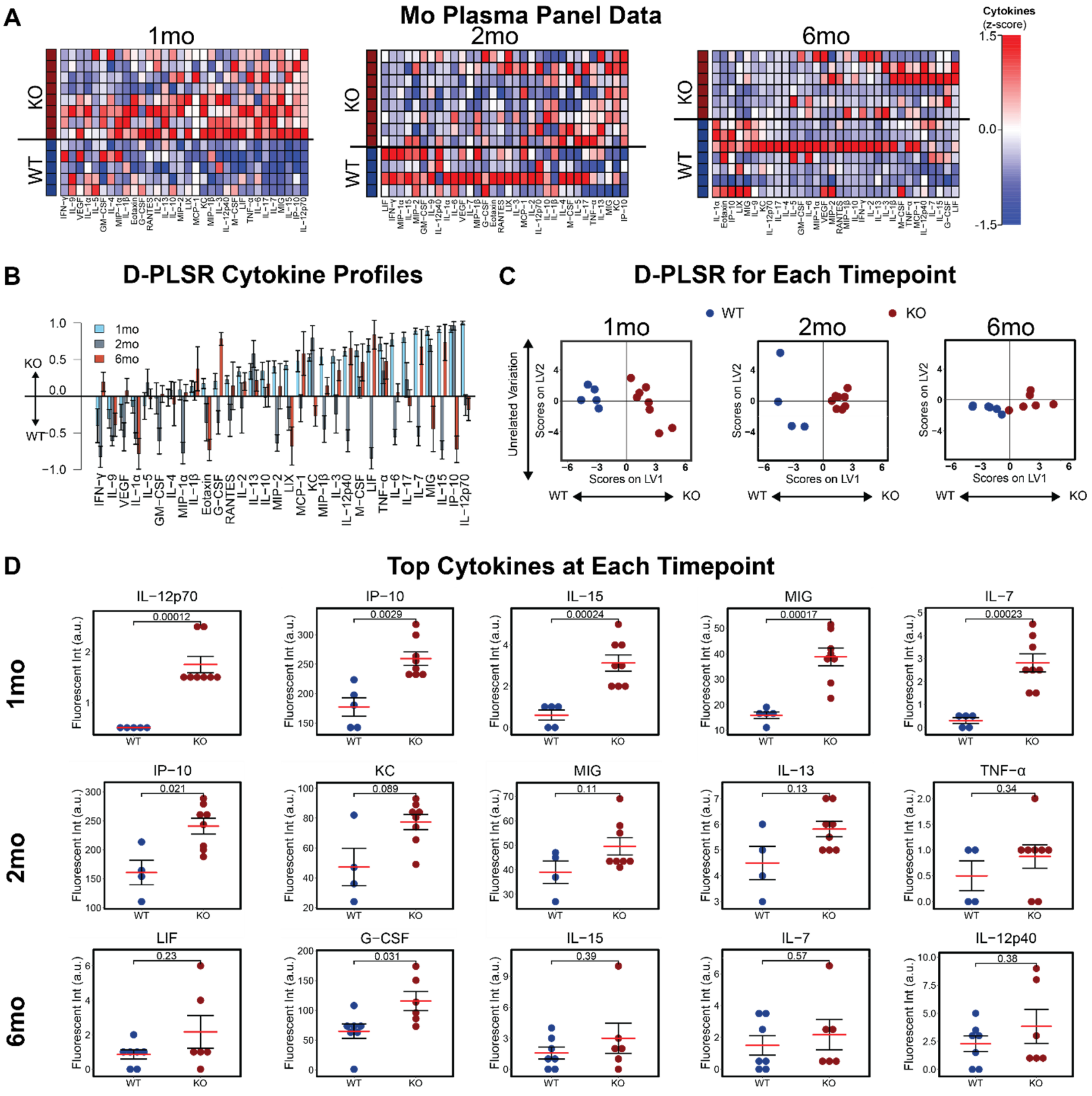


**Figure S2: Plasma cytokine signatures distinguish female *Mcoln1*$^{-/-}$ MLIV mice from wild-type controls.** (**A**) A panel of 32 cytokines quantified from blood plasma in WT (blue) and *Mcoln1*$^{-/-}$ KO (red) animals at 1, 2, and 6 months of age (each column is z-scored). (**B**) D-PLSR analysis at each timepoint reveals cytokine signatures associated with WT (negative) or KO (positive) mice (mean±SD in a LKOCV with K=1). (**C**) Scoring each sample from each time point based on its own LV1 profile in (B) separates WT mice to the left and KO mice to the right. (**D**) Univariate analysis of top cytokines from LV1 at each time point (mean±SEM, two-tailed t-test).

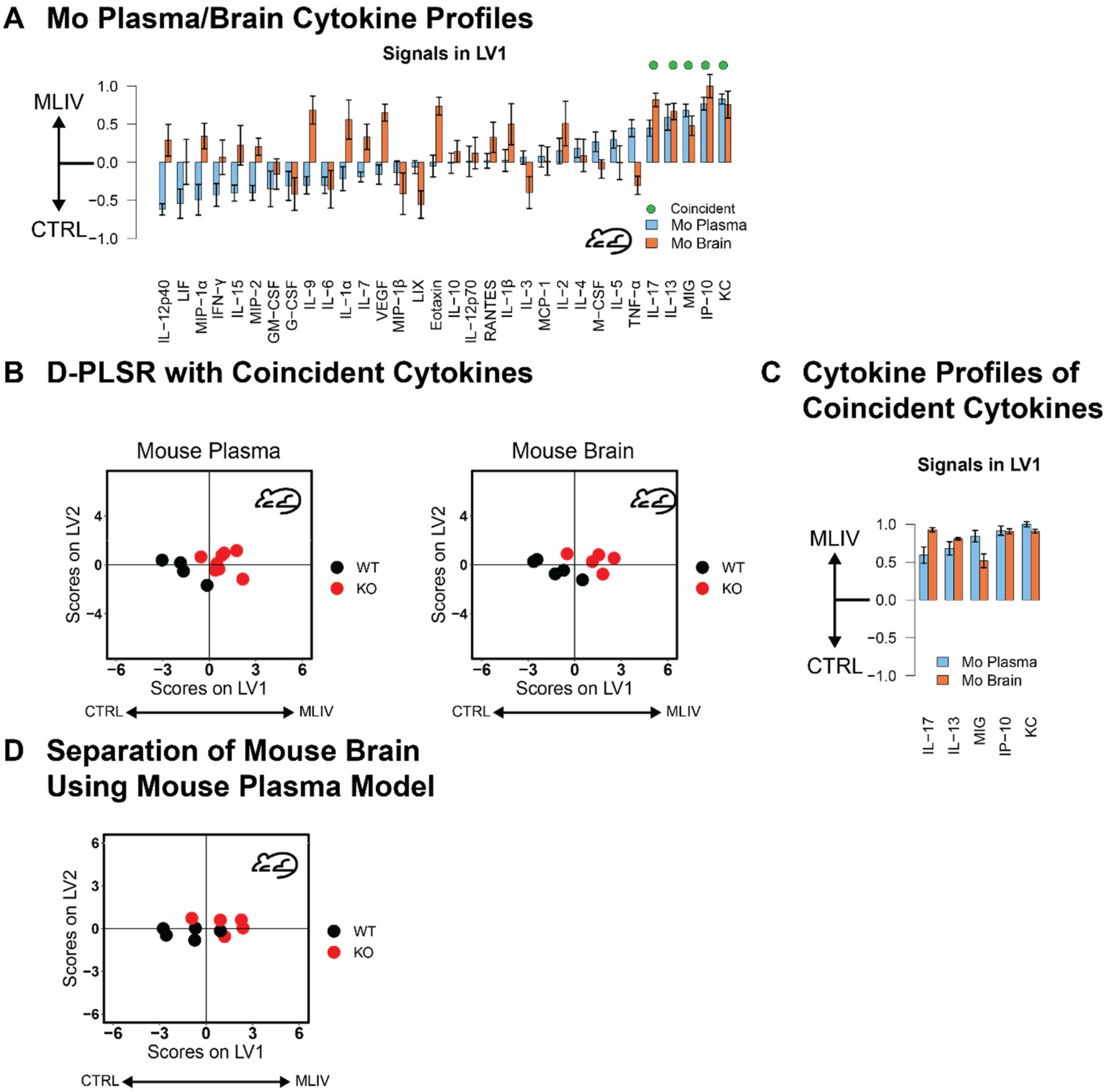


**Figure S3: Coincident cytokine signature distinguishes mouse MLIV plasma and mouse brain samples.** (**A**) LV1 cytokine profiles distinguishing mouse brain and human plasma generated based on 26 overlapping cytokines in human and mouse datasets. Of these, 5 cytokines were coincident in directionality to separate human plasma and mouse brain MLIV samples from controls. (**B**) D-PLSR analysis with 5 coincident cytokines separated both human plasma and mouse brain samples from controls along LV1, suggesting that this reduced cytokine signature is capable of separating control and MLIV samples from both huma plasma and mouse brain samples. (**C**) Reduced five cytokine LV1 cytokine profiles distinguishing MLIV and control cases for human plasma and mouse brain. (mean±SD in a LKOCV with K=1). (**D**) D-PLSR model generated based on 5 cytokines from human plasma separated wild-type mice to the left and KO mice to the right based on brain cytokine measurements.
